# Accelerated decline in white matter microstructure in subsequently impaired older adults and its relationship with cognitive decline

**DOI:** 10.1101/2020.09.23.20187450

**Authors:** Owen A. Williams, Andrea T. Shafer, Evian Perez Rivera, Yang An, Bennett A. Landman, Luigi Ferrucci, Susan M. Resnick

## Abstract

**INTRODUCTION:** Little is known about longitudinal decline in white matter (WM) microstructure and its associations with cognition in preclinical Alzheimer’s disease (AD).

**METHODS:** Longitudinal diffusion tensor imaging and neuropsychological testing from 50 older adults who subsequently developed mild cognitive impairment or dementia (subsequently impaired, SI) and 200 cognitively normal controls. Rates of WM decline were compared between groups using voxel-wise linear mixed-effects models. Associations between change in WM and cognition were examined.

**RESULTS:** SI had faster decline in fractional anisotropy (FA) in the right inferior fronto-occipital fasciculus (R.IFOF) and bilateral splenium of the corpus callosum. Decline in R.IFOF FA was related to decline in verbal memory, visuospatial ability, processing speed, and MMSE (p≤ 0.05). Decline in bilateral splenium FA was related to decline in verbal fluency, processing speed, and MMSE (p≤ 0.05).

**DISCUSSION:** Accelerated regional WM decline is characteristic of preclinical AD and related to domain specific cognitive decline.

## Introduction

Dementia is one of the leading causes of death and disability worldwide, and it is projected to affect over 151 million individuals by 2050 [1]. Alzheimer’s disease (AD) is the most common cause of dementia. AD is a progressive neurodegenerative disease with a long preclinical phase typically defined by the accumulation of beta amyloid (Aβ) and phosphorylated tau with subsequent acceleration of regional brain atrophy in the absence of clinical symptoms [2–5]. However, the nature of longitudinal changes in white matter (WM) microstructure in the preclinical phase of AD has been less well characterized.

Anti-amyloid clinical trials have failed to show that removal of Aβ is associated with improved cognitive outcomes [6]. It has been proposed that trials are not targeting the disease early enough [6, 7]. This has led to an increased focus on the asymptomatic preclinical stage of AD, with the hope that interventions administered before neuronal damage and symptom onset may be more effective [8, 9]. For such a strategy to be successful, sensitive biomarkers must be developed that can accurately detect signs of preclinical AD changes and provide surrogate markers of disease progression to assess the efficacy of disease-modifying treatments [10]. Thus, it is critical to define the earliest, and possibly subtle, changes in brain structure in preclinical AD to identify all viable biomarkers for AD.

Network-based theories of AD point to patterns of disconnection of spatially dispersed but functionally connected regions of the brain, e.g. the default mode network (DMN) [11, 12]. Functional networks are supported by the cerebral WM connecting the gray matter regions implicated in AD. It is therefore important to understand the spatial pattern of structural alterations of WM in the preclinical stages of AD. Diffusion tensor imaging (DTI) is an imaging technique that is sensitive to the microstructural properties of cerebral WM [13]. In the WM, axons, myelin sheaths and neurofilaments restrict both direction and magnitude of Brownian diffusion, leading to highly directional diffusion running in parallel with WM tracts. Fractional anisotropy (FA) is used to quantify the degree of anisotropic diffusion and mean diffusivity (MD) is used to quantify the magnitude of total water diffusion within brain tissue. Age-related decreases in FA and increases in MD have been widely reported from both cross-sectional and longitudinal studies and are thought to reflect microstructural damage that is associated with cognitive decline [14–18]. However, while preclinical AD is associated with declines in several cognitive domains including episodic memory, executive functioning, visuospatial functioning and processing speed [19, 20], few studies have examined the associations between changes in DTI metrics and changes in cognition in preclinical AD [21].

Compromised WM microstructure (lower FA, higher MD) in AD compared to cognitively normal (CN) controls has been reported in diffuse areas including the fornix, splenium of the corpus callosum, inferior fronto-occipital fasciculus, and uncinate fasciculus.[22–24] However, studies of alterations in WM microstructure during the preclinical phase have been less consistent. For example, in samples of individuals with autosomal dominant forms of early onset AD (EOAD), different regional findings or lack of associations have been reported across studies. In one study [25], FA was reduced in the fornix and orbitofrontal WM in preclinical EOAD patients compared with non-gene carrier relatives. However, other studies found no significant differences in DTI metrics between preclinical EOAD participants and non-carrier relatives [26, 27]. These studies all tend to have small samples of preclinical EOAD patients (typically less than 30) and are limited to cross-sectional analyses that do not assess within-individual changes in WM microstructure. Furthermore, EOAD represents a small percentage of cases of AD, and additional studies focusing on preclinical stages in late onset AD are needed.

The aim of the current study was to characterize the course of microstructural WM changes in preclinical AD and to interrogate their relationship with cognitive decline. To elucidate AD-related regional changes in WM microstructure prior to symptom onset, we investigated voxel-wise differences in changes in FA and MD in individuals who later developed mild cognitive impairment (MCI) or AD versus those who remained CN. We hypothesized that observed differences in rates of microstructural change would be identified in regions previously implicated in AD such as the cingulum, corpus callosum, and medial temporal lobe WM. Furthermore, we assessed the associations between localized rates of change in DTI metrics and rates of change in cognition in domains that show decline in AD.

## Materials and methods

### Participants

Participants were from the Baltimore Longitudinal Study of Aging (BLSA), a prospective study of physical and psychological aging that started in 1958 [28]. Participants were healthy at enrollment and those who were over time diagnosed with either MCI (n = 30) or dementia (n = 20) were matched using MatchIt [29] based on baseline age, sex, race, and follow-up time from first DTI in a 1:4 ratio with participants with normal cognitive status throughout follow-up. The BLSA study is approved by the local Institutional Review Boards, and participants provided written informed consent at each visit. Determination of cognitive status was performed through established procedures, please refer to Supplementary Material (SM) for more information. Table 1 provides the sample characteristics for both cognitive status groups (50 subsequently impaired (SI) and 200 CN participants). For the SI group, the last DTI scan included in the analysis preceded the date of symptom onset. There were no significant differences between cognitive groups for any of the matching criteria including baseline age, sex, race, and length of follow-up from first 3T DTI scan. There were also no significant differences in years of education, apolipoprotein E ɛ4 status, and vascular risk factors including hypertension, elevated cholesterol, diabetes mellitus, or obesity.

**Table 1.**
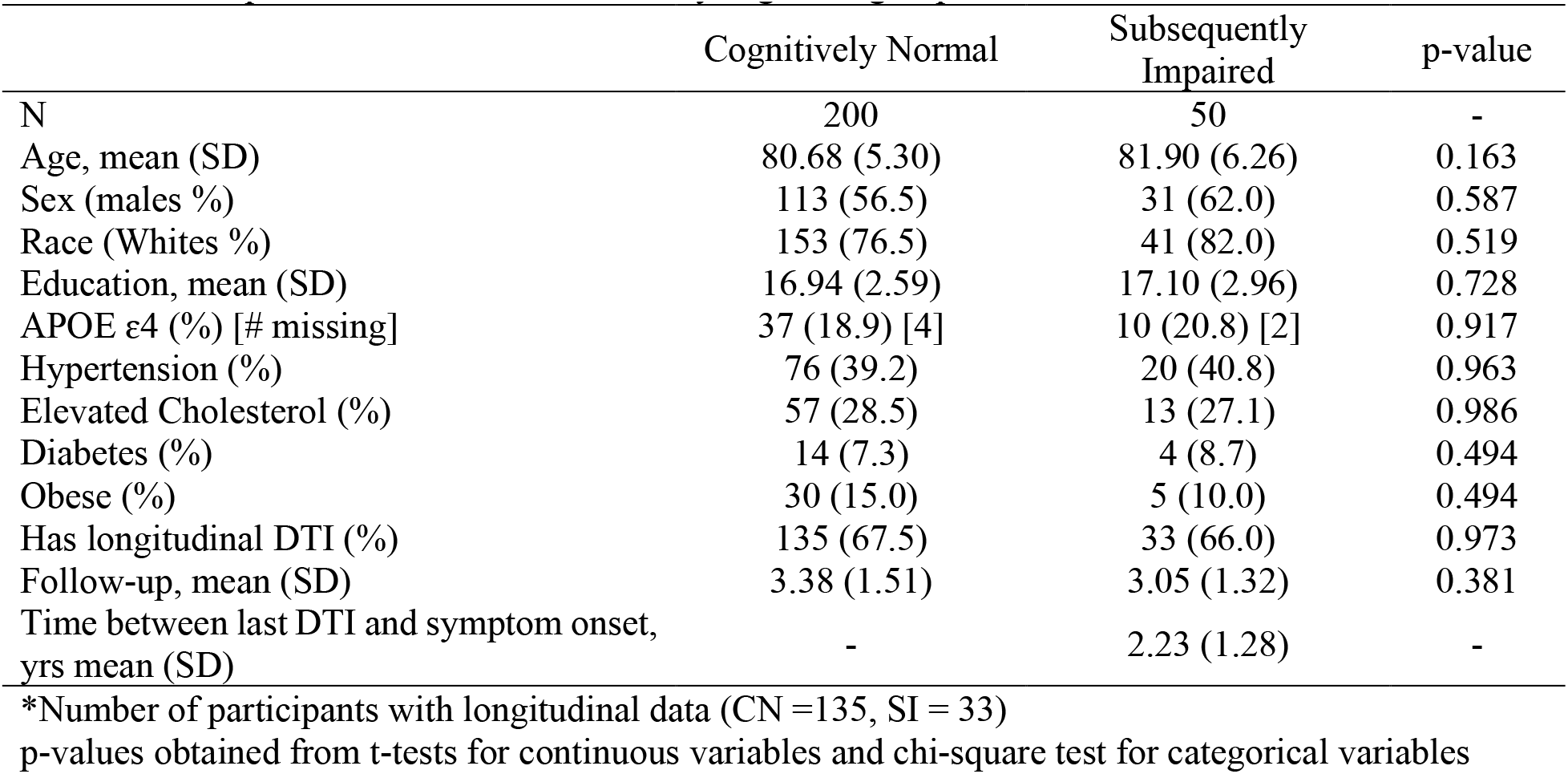
Participant characteristics stratified by cognitive groups

|  | Cognitively Normal | Subsequently Impaired | p-value |
| --- | --- | --- | --- |
| N | 200 | 50 | - |
| Age, mean (SD) | 80.68 (5.30) | 81.90 (6.26) | 0.163 |
| Sex (males %) | 113 (56.5) | 31 (62.0) | 0.587 |
| Race (Whites %) | 153 (76.5) | 41 (82.0) | 0.519 |
| Education, mean (SD) | 16.94 (2.59) | 17.10 (2.96) | 0.728 |
| APOE ε4 (%) [# missing] | 37 (18.9) [4] | 10 (20.8) [2] | 0.917 |
| Hypertension (%) | 76 (39.2) | 20 (40.8) | 0.963 |
| Elevated Cholesterol (%) | 57 (28.5) | 13 (27.1) | 0.986 |
| Diabetes (%) | 14 (7.3) | 4 (8.7) | 0.494 |
| Obese (%) | 30 (15.0) | 5 (10.0) | 0.494 |
| Has longitudinal DTI (%) | 135 (67.5) | 33 (66.0) | 0.973 |
| Follow-up, mean (SD) | 3.38 (1.51) | 3.05 (1.32) | 0.381 |
| Time between last DTI and symptom onset, yrs mean (SD) | - | 2.23 (1.28) | - |

\*Number of participants with longitudinal data (CN =135, SI = 33)
p-values obtained from t-tests for continuous variables and chi-square test for categorical variables

### DTI acquisition and preprocessing

MRI data were acquired on three different 3 Tesla Philips Achieva scanners (scanners 1 and 2 at the Kennedy Krieger Institute and scanner 3 at the National Institute on Aging). The DTI acquisition protocols are provided in SM. A general overview of the preprocessing steps is provided here, with more detailed information provided in SM. DTI data were corrected for physiological motion effects and eddy currents. Tensor fitting was carried out using FSL FDT using the ordinary least squares method. Resultant FA and MD maps were selected for analysis. Quality control methods for DTI data in the BLSA have been described previously[30, 31]. Baseline motion and motion over time did not differ between groups, see SM.

The tract-based spatial statistics (TBSS [32]) analysis utilized a BLSA-specific FA template was generated from FA images from 60 (30 men, 30 women) cognitively normal BLSA participants with a mean age of 69.9 years (range 60-80 years) using advanced normalization tools (ANTS) MultivariateTemplateConstruction2 [33]. TBSS was used to generate a WM skeleton to facilitate voxel-wise analysis in WM tracts common to all participants. See SM for details of the processing pipeline.

### Cognitive measures and domains

Composite scores were calculated for verbal memory, executive function, attention, visuospatial ability, verbal fluency, and processing speed. The BLSA neuropsychological test battery has been described previously [31, 34] and details are provided in SM, Table S1. We also examined performance on a measure of mental status, the mini-mental state examination (MMSE) [35].

### Statistical analyses Differences in longitudinal WM microstructural trajectories due to subsequent impairment

Voxel-wise analysis was performed in MATLAB (Natick, Massachusetts: The MathWorks Inc.). To estimate group differences in longitudinal change in WM microstructure, linear mixed-effects models were implemented using fitlme (https://www.mathworks.com/help/stats/fitlme.html) with FA or MD as the dependent variable. The analysis was restricted to voxels in the mean FA skeleton image. Intercept and time (follow-up time in years from first 3T DTI scan) were entered as random effects with unstructured covariance. Fixed effects included cognitive group (SI/preclinical = 1, cognitively stable = 0), baseline age (mean centered), sex, race, scanner, baseline motion (mean FD), and 2-way interactions of cognitive group, baseline age, sex, and race with time. To control for the effect of multiple comparisons, contrasts of interest (the main effect of cognitive group and cognitive group*time interaction) were examined using a p-value of less than 0.005 and cluster size of 30 voxels or more and also using a more conservative approach, false discovery rate (FDR) [36]. For each cluster that met these requirements, the coordinates of the peak voxel (defined as highest t-value) were used to define the location of the cluster by comparison with the Johns Hopkins University ICBM-DTI-81 WM labels atlas [37]. While the primary focus of this study was to assess group differences in *longitudinal* change in DTI metrics, we also report baseline results in SM.

### Relationship between change in FA and change in cognition

We first used linear mixed models to examine cognitive change in the SI relative to CN groups in this subsample with concurrent cognitive and DTI data. The same linear mixed-effects models were used to estimate group differences in longitudinal change as were used for the DTI data minus the fixed effect for scanner (R 3.3.2, nlme 3.1-139). Next, we assessed the amount of between-person variability in preclinical longitudinal change explained by the effect of interest (subsequent cognitive group) for each outcome (cognition and DTI). The amount of variance explained (*R^2^*) in the change trajectory accounted for by cognitive group is a measure of effect-size used in multilevel models [38, 39] and can provide an estimate of how sensitive the outcome is in detecting differences in change over time due to preclinical disease, see SM for additional information.

Next, we examined the relationship between change in FA and change in cognition, focusing on the three larger uncorrected FA clusters that survived FDR. Partial correlations between estimated slopes controlling for age, sex, and race were used to assess the relationship between change in mean FA and change in cognition, see SM for more information.

## Results

### Faster rates of WM microstructural change in the splenium and inferior fronto-occipital fasciculus in those with subsequent impairment

Results from the LME models for the cognitive group*time interaction revealed 10 clusters where FA declined at faster rates in the SI group compared to the CN group over time (Table 2). The largest areas of decline were in the splenium of the corpus callosum and in the inferior fronto-occipital fasciculus, see Figure 2. Clusters in these areas survived FDR correction for multiple comparisons, but this correction resulted in smaller, separate clusters. For example, the largest cluster (right splenium of the corpus callosum, 728 voxels) became three smaller clusters of 124 voxels, 107 voxels and 67 voxels (Table 2). Other areas showing faster declines in FA were the superior cerebellar peduncle, postcentral gyrus WM, middle/lateral occipital gyrus WM, cingulate gyrus WM, genu of the corpus callosum, and posterior corona radiata. However, these clusters did not survive FDR correction.

**Table 2.**
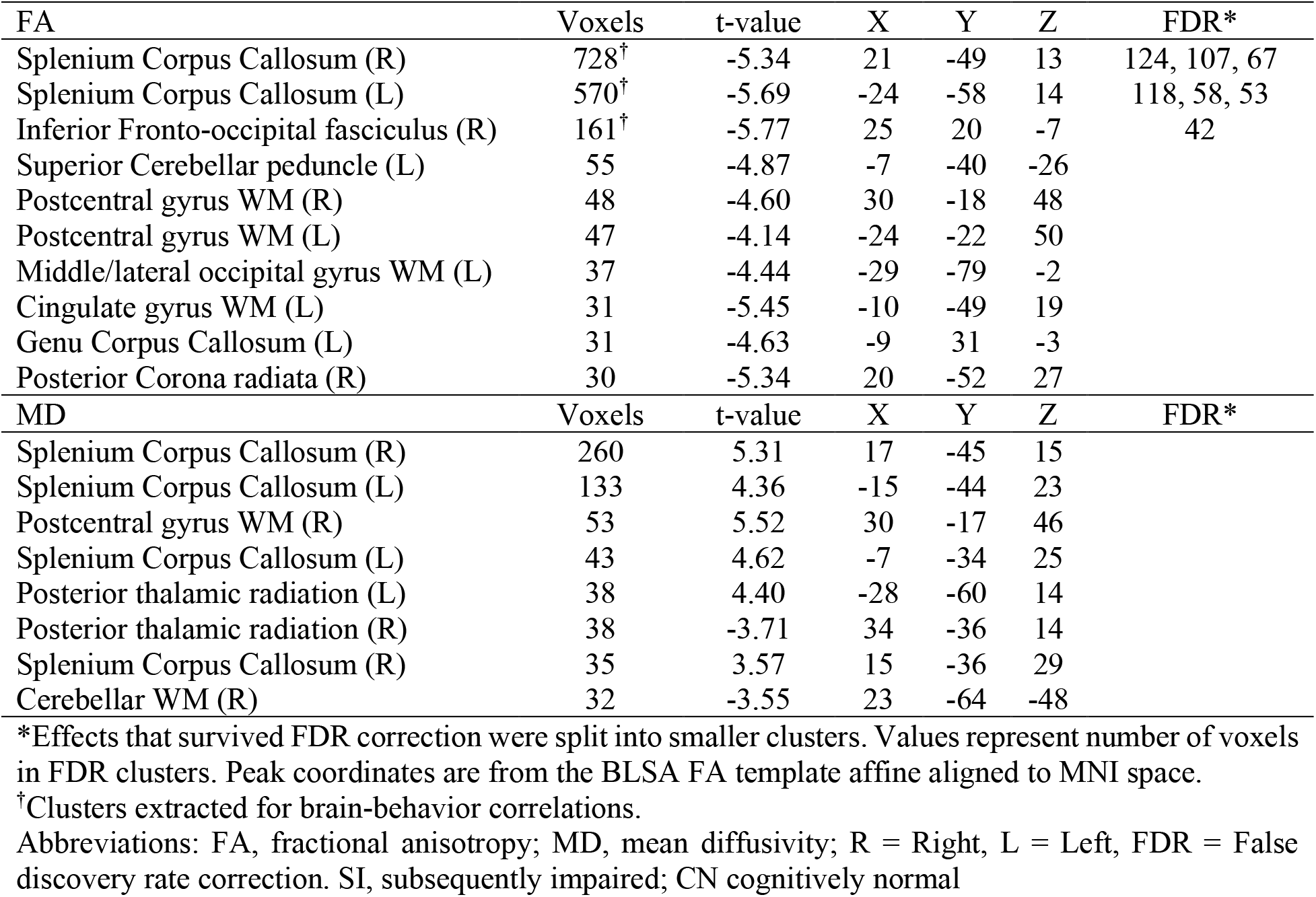
White matter clusters showing differences in the rates of change in FA and MD between SI and CN groups. The peak t-value is reported for each cluster.

| FA | Voxels | t-value | X | Y | Z | FDR* |
| --- | --- | --- | --- | --- | --- | --- |
| Splenium Corpus Callosum (R) | 728<sup>†</sup> | -5.34 | 21 | -49 | 13 | 124, 107, 67 |
| Splenium Corpus Callosum (L) | 570<sup>†</sup> | -5.69 | -24 | -58 | 14 | 118, 58, 53 |
| Inferior Fronto-occipital fasciculus (R) | 161<sup>†</sup> | -5.77 | 25 | 20 | -7 | 42 |
| Superior Cerebellar peduncle (L) | 55 | -4.87 | -7 | -40 | -26 |  |
| Postcentral gyrus WM (R) | 48 | -4.60 | 30 | -18 | 48 |  |
| Postcentral gyrus WM (L) | 47 | -4.14 | -24 | -22 | 50 |  |
| Middle/lateral occipital gyrus WM (L) | 37 | -4.44 | -29 | -79 | -2 |  |
| Cingulate gyrus WM (L) | 31 | -5.45 | -10 | -49 | 19 |  |
| Genu Corpus Callosum (L) | 31 | -4.63 | -9 | 31 | -3 |  |
| Posterior Corona radiata (R) | 30 | -5.34 | 20 | -52 | 27 |  |
| MD | Voxels | t-value | X | Y | Z | FDR* |
| Splenium Corpus Callosum (R) | 260 | 5.31 | 17 | -45 | 15 |  |
| Splenium Corpus Callosum (L) | 133 | 4.36 | -15 | -44 | 23 |  |
| Postcentral gyrus WM (R) | 53 | 5.52 | 30 | -17 | 46 |  |
| Splenium Corpus Callosum (L) | 43 | 4.62 | -7 | -34 | 25 |  |
| Posterior thalamic radiation (L) | 38 | 4.40 | -28 | -60 | 14 |  |
| Posterior thalamic radiation (R) | 38 | -3.71 | 34 | -36 | 14 |  |
| Splenium Corpus Callosum (R) | 35 | 3.57 | 15 | -36 | 29 |  |
| Cerebellar WM (R) | 32 | -3.55 | 23 | -64 | -48 |  |

\*Effects that survived FDR correction were split into smaller clusters. Values represent number of voxels in FDR clusters. Peak coordinates are from the BLSA FA template affine aligned to MNI space.
<sup>†</sup>Clusters extracted for brain-behavior correlations.
Abbreviations: FA, fractional anisotropy; MD, mean diffusivity; R = Right, L = Left, FDR = False discovery rate correction. SI, subsequently impaired; CN cognitively normal

**Figure 1.**
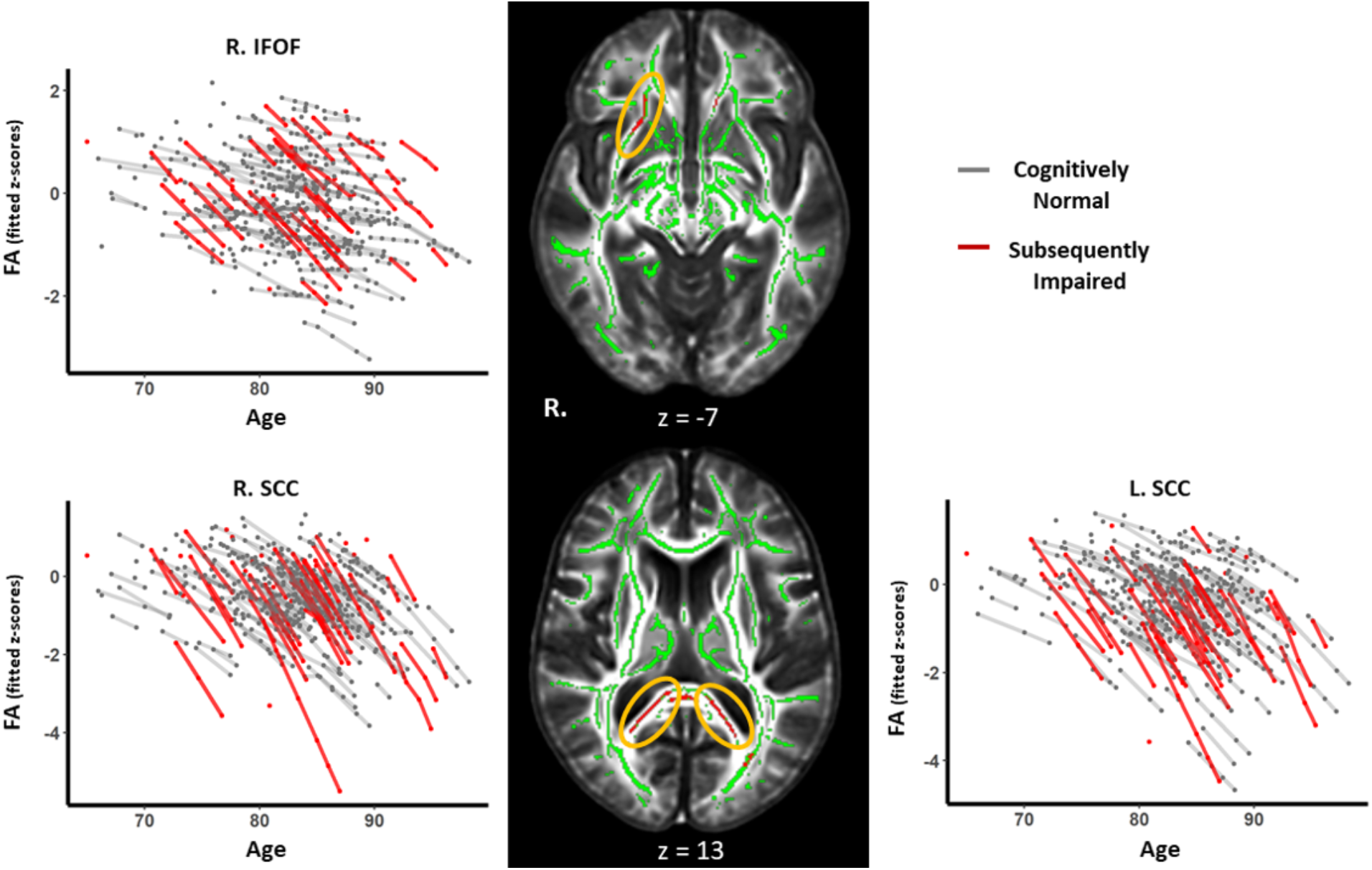
White matter clusters showing accelerated microstructural decline in the SI group. Line graphs show modelled trajectories of decline in FA for clusters in the right inferior fronto-occipital fasciculus, right splenium of the corpus callosum, and left splenium of the corpus callosum. The black panel shows axial slices where clusters with accelerated FA decline were found for the SI group in the right inferior frontooccipital fasciculus (top) and bilateral splenium of the corpus callosum (bottom) highlighted in red and indicated by the yellow ellipsoids.

**Figure 2.**
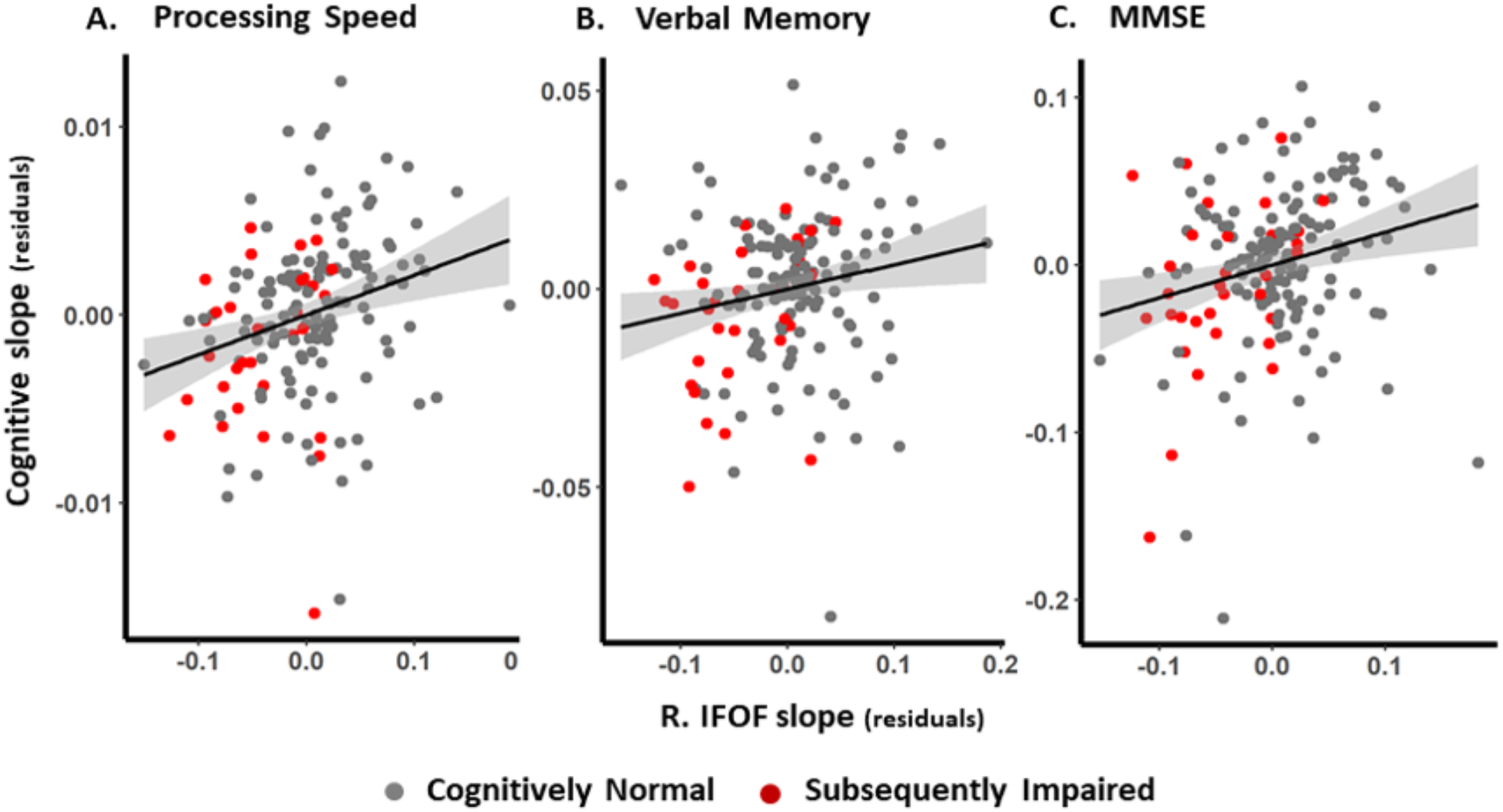
Associations bewteen change in FA and change in cognition. Figure shows the relationship between the rate of change in the right inferior fronto-occipital fasciculus and the rate of change in processing speed (panel A), verbal memory (panel B), and MMSE (panel C). The effects of age, sex, and race were removed from the estimated slopes for FA and cognition. Shaded areas indicate 95% confidience intervals.

Six clusters were found to show faster increases in MD over time for the SI relative to the CN group and two clusters showed faster increases in MD in the CN vs. SI group (Table 2). Of the six clusters showing faster increase in MD for the SI group, four were in the splenium of the corpus callosum and the remaining two clusters were in the R. postcentral gyrus and L. posterior thalamic radiation. The two clusters with slower increases in MD in the SI compared to the CN group were in the R. posterior thalamic radiation and R. cerebellar WM. None of the clusters showing longitudinal change in MD survived FDR correction.

### Correlations between change in FA and change in cognition

Linear mixed-effects models controlling for age, sex, and race were performed to cognitively characterize this DTI subsample of the BLSA. Significantly faster rates of cognitive decline in SI compared with CN participants were observed for memory, executive function, and processing speed (see Table 3 and Supplementary Material, Figure S1), but not for spatial ability or attention. Effect sizes for the group differences in rates of longitudinal change were estimated using an *R^2^* statistic for multilevel models in which the amount of residual variance in between-person slopes captured by cognitive status was calculated for cognitive and DTI measures. These showed that the cognitive status grouping explained more between-person variation in the longitudinal trajectories of FA (L.SCC, 76%; R. SCC, 81%; R. IFOF, 61%) when compared to the longitudinal trajectories of the cognitive measures. Within the cognitive domains examined, longitudinal trajectories for processing speed (42%) and memory (25%) were the most sensitive to subsequent cognitive status, see Table 3.

**Table 3.** Differences between SI and CN groups in longitudinal rates of change in cognition and the three main FA clusters showing differences. The linear mixed-effects model controlled for inter-individual differences in age, sex, and race. The results reported below for the FA clusters were derived from z-scored, baseline anchored mean raw FA values extracted from the clusters. Here, the models are identical for the cognitive and FA data. Effect size for differences in longitudinal trajectories between SI and CN groups was determined using a *R^2^* statistic for multilevel models in which the residual variance in between-person slopes captured by including the effect of subsequent impairment was estimated.

| **Cognitive Domain** | **Beta** | **P-value** | **Effect Size** |
| --- | --- | --- | --- |
| Verbal Memory | **-0.075\*** | **0.044** | 24.93 |
| Executive Function | **-0.081\*** | **0.039** | 18.53 |
| Attention | -0.059 | 0.079 | 7.21 |
| Verbal Fluency | -0.056 | 0.079 | 11.06 |
| Visuospatial Ability | -0.02 | 0.530 | -11.06 |
| Processing Speed | **-0.119†** | **< 0.001** | 41.73 |
| MMSE | -.099 | 0.096 | 18.86 |
| **WM ROI<sup>+</sup>** | **Beta** | **P-value** |  |
| Right SCC | **-0.284†** | **< 0.001** | 84 |
| Left SCC | **-0.254†** | **< 0.001** | 75.59 |
| Right IFOF | **-0.175†** | **< 0.001** | 60.78 |

\* significant at $p \leq 0.05$ .
† significant at $p \leq 0.01$ .
<sup>+</sup> All datapoints (subjects and visits) were utilized for the WM ROIs, however, similar results were obtained in analyses based on matched sample sizes between models.
Abbreviations: MMSE, mini-mental state examination; SCC, splenium of corpus callosum; IFOF, Inferior fronto-occipital fasciculus; SI, subsequently impaired; CN cognitively normal.

For the three cognitive domains that showed significantly faster decline in SI participants (processing speed, memory, and executive function), decline in processing speed was associated with decline in FA across all three regions showing greater FA decreases in the SI group (L. SCC, ρ = 0.169, p = 0.034; R. SCC, ρ = 0.180, p = 0.023; R. IFOF, ρ = 0.265, p = 0.001, see Figure 3A). Decline in episodic memory was associated with FA decline in the right IFOF (ρ = 0.182, p = 0.020, see Figure 3B), whereas no associations were found between decline in executive function and FA in the clusters examined, see Table 4. Of the cognitive domains and MMSE that did not show significantly faster decline for SI participants, decline in MMSE was linked to decline in FA in all clusters, see Figure 3C and Table 4. Decline in verbal fluency was associated with FA decline in bilateral SCC and decline in visuospatial ability with FA decline in the R. IFOF.

**Table 4.** Associations between change in FA and change in cognition. Associations were determined by partial correlations controlling for age, sex, and race. Partial correlations were performed between slopes for subjects with longitudinal data.

|  | Left Splenium CC |  |  | Right Splenium CC |  |  | Right IFOF |  |  |
| --- | --- | --- | --- | --- | --- | --- | --- | --- | --- |
|  | Pearson's r | P-value | 95% CI | Pearson's r | P-value | 95% CI | Pearson's r | P-value | 95% CI |
| VM | 0.106 | 0.183 | -0.05,0.26 | 0.126 | 0.111 | -0.03,0.28 | **0.182** | **0.021\*** | **0.03,0.33** |
| EF | 0.043 | 0.606 | -0.12,0.19 | -0.038 | 0.649 | -0.2,0.11 | 0.045 | 0.590 | -0.12,0.2 |
| ATT | -0.002 | 0.98 | -0.16,0.15 | 0.01 | 0.87 | -0.14,0.17 | -0.05 | 0.52 | -0.2,0.1 |
| VF | **0.19** | **0.02\*** | **0.04,0.33** | **0.23** | **0.003\*** | **0.08,0.37** | 0.05 | 0.52 | -0.1,0.2 |
| VSA | 0.069 | 0.386 | -0.09,0.22 | -0.01 | 0.9 | -0.16,0.15 | **0.184** | **0.019\*** | **0.03,0.33** |
| PS | **0.167** | **0.038\*** | **0.01,0.32** | **0.179** | **0.025\*** | **0.03,0.33** | **0.266** | **0.005\*\*** | **0.11,0.4** |
| MMSE | **0.16** | **0.05\*** | **0.003,0.3** | **0.185** | **0.02\*** | **0.03,0.33** | **0.23** | **0.004\*** | **0.08,0.37** |

\* significant at $p \leq 0.05$ .
\*\* significant at $p \leq 0.01$ .
Abbreviations: VM, verbal memory; EF, executive function; ATT, attention; VF, verbal fluency; VSA, visuospatial ability; PS, processing speed; SCC, splenium of corpus callosum; IFOF, Inferior fronto-occipital fasciculus.

## Discussion

Using a case-control subset of the BLSA, we found that faster rates of WM microstructural degeneration are detectable before clinical symptoms of MCI or AD are apparent. Longitudinal voxel-wise analyses identified several clusters (bilateral splenium of corpus callosum and right inferior frontooccipital fasciculus) where FA declined faster in a sample of subsequently impaired individuals compared to a control group who remained CN. While several overlapping clusters were found where MD was increasing faster in the SI group compared to controls, these clusters did not remain significant after FDR correction. Furthermore, rates of decline in WM microstructure were associated with rates of decline in cognition in domains that did and did not show significant differences in rates of decline in cognitive status groups.

The largest clusters to show faster decline in FA in the SI compared to CN group, and survive FDR correction, were in the left and right splenium of the corpus callosum. This bilateral pattern of accelerated change suggests that the splenium may be susceptible to microstructural decline in individuals on a trajectory towards MCI/AD. Our results extend prior reports of atrophy and reduced microstructure of the splenium of the corpus callosum in MCI/AD participants compared to CN participants [40] and in at risk APOE e4 carriers who show faster decline in FA in this area.[31] Here we have shown that such changes can be detected in the preclinical stages, before onset of clinical symptoms. The third largest cluster was in the inferior fronto-occipital fasciculus. The microstructure of the IFOF has been shown to decline faster in AD patients compared to CN controls [23] and show cross-sectional differences in DTI metrics between preclinical EOAD participants and controls. [25]. The splenium of the corpus callosum contains commissural fibers that connect association regions in the temporal and parietal lobes, including the precuneus[41], while the IFOF contains fibers connecting frontal and occipital lobes. Evidence suggests that the precuneus and frontal lobe areas are among the earliest regions to accumulate Aβ.[42] Therefore it is possible the microstructural decline is related to pathological changes occurring in these regions early in the preclinical stage of AD.

We found no evidence of faster microstructural decline in limbic structures, including the fornix. This contrasts with Ringman et al. [25] who found cross-sectional evidence of lower FA in the fornix in comparing preclinical EOAD to non-gene carrier family members. This difference may be due to Ringman et al.’s use of small ROIs (two voxels) placed in each subject’s native space FA maps rather than a TBSS approach. In another cross-sectional study of preclinical EOAD participants compared to CN controls using TBSS, no difference in DTI markers in the fornix was reported. [43]

Cognitively characterizing this DTI subsample of the BLSA confirmed that the SI group had similar preclinical cognitive profiles to other preclinical samples [19, 44]. Cognitive trajectories for three domains (verbal memory, executive function, and processing speed) showed faster decline in the SI group. The pattern for all cognitive outcomes examined, apart from visuospatial ability, was similar with trends toward greater decline the SI group. The exception of visuospatial ability is interesting as our recent publication of change-points in AD in a larger BLSA preclinical sample found the earliest change-point in cognition for these participants was visuospatial ability [20]. It is possible that the relatively short follow-up in the current sample limited our ability to detect group differences. Furthermore, the inclusion of individuals with MCI in the SI sample in the present study was necessary to increase sample size due to the more recent introduction of DTI. The heterogeneity in the SI sample due to the inclusion of both MCI and AD endpoints, and heterogeneity in cognitive trajectories, may explain the lack of significant differences in visuospatial trajectories over time in this BLSA subsample.

Comparing the effect size (*R^2^*) for group membership (proportion of explained variance in rates of change by group membership), we found FA to yield considerably higher values compared to the cognitive outcomes. These findings are not surprising as the cognitive data are more variable over time due to a greater number of factors influencing performance. However, these findings suggest that regional changes over time in DTI markers may provide a more robust marker of changes during preclinical AD.

Associations between change in FA and change in cognition were observed for five domains. Of the three domains that showed significant differences in decline by group, faster decline in FA in the R IFOF was associated with the faster of decline in verbal memory, and faster decline in FA in all three clusters was related to faster decline in processing speed. Additionally, faster FA decline in all three clusters was related to MMSE, while the splenium clusters were linked to declines in verbal fluency and the R IFOF to visuospatial ability. These results extend previous findings that regional differences in frontal lobe WM microstructure between AD and controls was also correlated with memory performance.[44] While splenium FA decline was not related to verbal memory, it was related to verbal fluency, processing speed, and MMSE. These results are consistent with a previous cross-sectional report showing FA in the splenium is associated with poorer cognition in MCI/AD patients as measured by the clinical dementia rating scale.[45] The finding that processing speed was related to FA decline in all three clusters is perhaps less surprising given previous reports of significant associations between WM microstructure in these areas and processing speed in CN older adults.[46]

Results from this study should be considered in the context of several limitations. BLSA participants in this sample are highly educated, mostly white, and relatively old, with a mean baseline age of 80 years, which may limit generalizability. Furthermore, the follow-up period was relatively short, and while most participants had multiple DTI and cognitive assessments, some only contributed cross-sectional data. However, the sample size and longitudinal power are still improvements on previous studies of preclinical changes in DTI. The SI sample included participants, who were cognitively normal at the time of imaging but had research diagnoses of either MCI or AD as their final cognitive status at the time of analysis. While MCI is typically the prodromal stage of AD [47], it is possible that some of MCI participants will remain stable or develop non-AD pathologies. This heterogeneity may increase variance in the SI group, leading to an underestimation of the extent of localized WM microstructural damage in preclinical AD. Future work with larger samples of AD cases may address this issue. The study benefits from statistical matching of SI participants to CN participants with similar demographic features, allowing for higher confidence that the differences found between groups are indeed a result of the group differences in disease trajectories. However, we cannot exclude the possibility that some individuals in the CN group will develop cognitive impairment later, and it is possible that more robust differences may be found in samples where AD pathology is confirmed postmortem.

In this study, longitudinal DTI data were used to characterize voxel-wise patterns of accelerated decline in WM microstructure and its relationship to cognition during the preclinical phase of MCI/AD. Group membership explained more variance in rates of change in regional brain microstructure than it did in cognitive decline, suggesting DTI may be a powerful indicator of future risk of developing MCI/AD. In order to find the optimal imaging markers of preclinical AD, future work should focus on elucidating the joint associations and temporal relationships between change in regional DTI metrics and changes in other markers of AD such as atrophy and Aβ accumulation in the preclinical stage.

## Data Availability

Data from the BLSA are available on request from the BLSA website (http://blsa.nih.gov). All requests are reviewed by the BLSA Data Sharing Proposal Review Committee and are also subject to approval from the NIH institutional review board.

## Acknowledgements

We thank the staff of the BLSA and the NIA 3T MRI facility for their assistance and the BLSA participants for their dedication to this study.

## Funding

This research was supported by the Intramural Research Program of the National Institutes of Health, National Institute on Aging.

## Competing Interests

Declaration of interests: None.

## Notes

### Competing Interest Statement

The authors have declared no competing interest.

### Clinical Trial

NCT00233272

### Funding Statement

This research was supported entirely by the Intramural Research Program of the National Institutes of Health, National Institute on Aging.

### Author Declarations

Research protocols were approved by by the National Institute of Environmental Health Sciences (NIEHS) IRB which provided ethical approval. All participants gave written informed consent at each visit.

